# Assessing Language Difficulties in Health Facilities in Malawi

**DOI:** 10.1101/2024.11.06.24316625

**Authors:** Amelia Taylor, Paul Kazembe

## Abstract

Language barriers in healthcare, prevalent globally, often lead to miscommunication between professionals and patients, reducing the quality of care. Recognition, training, and formal study of these barriers during medical communication in African countries is severely limited. Our study focused on language barriers in healthcare facilities in Zomba, an important district in Malawi where several local languages are spoken. We employed a qualitative approach and conducted a questionnaire-based study. Data was gathered from 79 healthcare professionals and 312 patients. Our findings show that a lack of language preparation during health training and the absence of suitable resources in local languages, such as dictionaries, result in healthcare professionals struggling to communicate with patients in both English and local languages about medical issues. Patients also face difficulties in fully conveying their symptoms and understanding their diagnoses or treatments due to language barriers. We conclude that language training and health dictionaries in both English and local languages can help address some of these observed language barriers.

## 1. Background

Language barriers in healthcare exist in many countries across the world and have been shown to cause miscommunication between healthcare professionals and patients that lead to decreasing quality of healthcare [1], [2], [3], [4]. Several studies exist in developed countries such as the USA and Europe where large scale international migration has been the main source of language barriers [5], [6], [7]. On the African continent, South Africa has the largest number of studies that investigated language barriers in healthcare. Some focussed on the impact of language in translating informed consent when collecting health data [8], others focussed on language barriers that arose when staff who communicated mainly in English or Afrikaans, interacted with patients who spoke Xhosa. The language barriers reported were considerable and persistent and had multiple ramifications, from negatively influencing the attitudes of patients and staff towards each other [8] to reducing compliance and affecting emergency care [9]. A recent study in Ethiopia where Amharic is the official language, and many other local languages are spoken, showed evidence of similar language discordance between patients and healthcare providers [10].

Africa has many local languages that are not confined to territorial borders but followed long established migrations across the continent. For example, Chichewa is a Bantu language used in Malawi and in several other countries in Southern Africa [11]. Due to colonialism and widespread multilingualism, the primary language of healthcare and health education is not the language used in general communication by local communities. European languages are the official or the lingua franca in many countries. In Malawi, English is the official language and the main language for education, publishing, government, administration, law, finance and so on [12], [13]. Chichewa and other local languages are the preferred means for communication and for social cohesion. This language dichotomy underpins the existence of separate worlds in which English and local languages coexist to play complementary and non-overlapping roles. In these worlds inequalities exist in terms of accessing knowledge and services, including in healthcare. Healthcare professionals receive their training solely in English but must work and communicate with patients in local languages.

Despite the importance of language in health, we did not find other studies conducted in healthcare organisations in Malawi that focus on language barriers for health delivery. Our research seeks to understand the challenges language differences creates for health in Malawi. The insights from our study can inform policy and energise subsequent research into these issues and provide solutions.

## 2. Aims and Objectives of this research

We examine the use of language in patient-healthcare practitioner consultations in Malawi. We formulate the following research hypotheses. Healthcare professionals have difficulties in communicating with patients in English and in local languages about medical issues. Patients are not able to fully communicate their symptoms to healthcare professionals and they do not understand the diagnosis or the treatment their receive due to language barriers. In our research difficulties in communication cover those related to understanding spoken language and difficulties with the social aspects of communication and interaction. To test these two hypotheses, it is important to unpack their meaning.

*Proposition 1*. There is a lack of language preparation during health training and of suitable resources in local languages such as medical dictionaries.

*Proposition 2*. Healthcare professionals have difficulties in communicating with patients in English and in local languages about medical issues.

*Proposition 3*. Patients are not able to fully communicate their symptoms to healthcare professionals and they do not understand the diagnosis or the treatment their receive due to language barriers.

Together, these hypothetical propositions (which together form the hypothesis) consist of the following four main variables:

- *Independent variables*: language training, institutional requirements for language use in the workplace in taking notes, communicating diagnosis and treatment to patients, writing reports, the ability of medical professionals to use medical terms in local languages, and the ability of patients to express their symptoms and conditions in local languages or English.
- *Dependent variable*: language barriers in effective communication between healthcare professionals and patients.

The component propositions (or assumptions) of our hypothesis will be subjected to the data collected from fieldwork. To so that we break down proposition 1 into sub-components based on our local knowledge of the topic and from our background investigation.

A. Teaching and learning for healthcare professionals are done solely in English.
B. Communication in the workplace among healthcare professionals when discussing patient conditions and medical history takes place in English with mixed use of local languages.
C. Medical or hospital notes and reports are written only in English.
D. Healthcare professionals do not have access to medical dictionaries.
For proposition 2, we will collect data about the following assumptions:
E. Notes, diagnoses, and medication recorded in the patients’ health passports are in English.
F. Communication with patients requires the use of the patient’s language, which is in many cases different than that of the healthcare professional.
G. Healthcare professionals do not often explain medical terms to their patients.
For proposition 3 we will collect data about the following assumptions:
H. Healthcare professionals use notes written in previous visits in the health passport of the patient.
I. Patients rarely know what is written in their health passport.
J. Both healthcare professionals and patients struggle to express medical terminology in local languages.

### 2.1. Ethical Approval

The study was approved by the Research Committee of the District Health Office in Zomba that issued a letter of approval and a list of all facilities that came under its jurisdiction in the Zomba district. The letter enabled us to obtain separate approvals from health facilities to speak to healthcare professionals and patients on our visits. All participants were given a written explanation of the aim of the study and gave their consent verbally. We did not collect names of people; hence the consent of healthcare professionals came in the form of returning the completed questionnaires to the researcher. Patients gave verbal consent. Participants were assigned codes that were used throughout the data collection and management process to maintain confidentiality.

**Figure 1.**
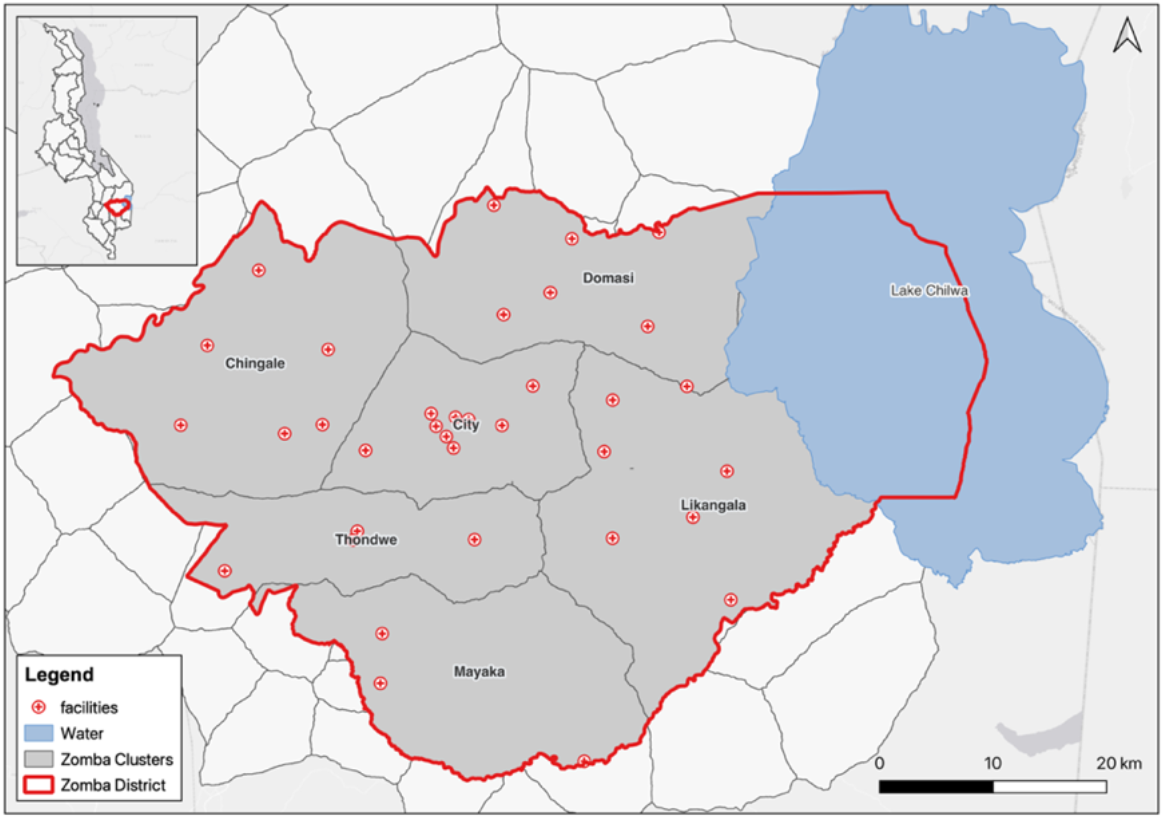
The study area in Zomba district showing facilities split per clusters

## 3. Methodology

We adopt a mixed qualitative approach guided by objectivism. The quantitative data resulting from the questionnaire was nominal and was analysed using descriptive statistics. We adopted the approach described in [14] by starting with a research hypothesis that informed our data collection. To validate the hypothesis, we employed a visual diagram-based approach for data analysis based on cause-and-effect evidence to determines the relationship between variables.

### 3.1. Study Site

The study was conducted in Zomba district, where Chichewa, Chiyao, and Chilomwe are spoken. Zomba city, the first capital of Malawi, is one of the four urban centers in the southern region and has a higher population density than the regional average. The district has 40 health facilities, mostly government-run, organized into six zones: Chingale, City, Domasi, Likangala, Mayaka, and Thondwe. A 2009 socio-economic report [15] highlighted acute shortages of healthcare staff, isolated health centers lacking electricity, running water, or telephones, and poor road conditions making transport difficult. By 2009, only four facilities had doctors, 15 had no medical assistant, and there was only one laboratory assistant for the district. The doctor-to-population ratio was 1:145,791, about three times higher than the Ministry of Health’s recommendation. We covered all facilities in Chingale, City, and Domasi zones, and one facility in Likangala. Mayaka and Thondwe were not covered due to their distance and our assessment that additional data would not significantly enhance the study.

### 3.2. Data collection through questionnaires

Data was collected through face-to-face questionnaires with healthcare professionals and patients. The authors developed this questionnaire due to the lack of existing validated tools for assessing language barriers in health in Malawi. At each clinic, we met the officer in charge at the start of the day, presented our approval letter from the District Health Office in Zomba, along with the questionnaires and agreement forms. After reviewing these documents, sometimes requiring a one-day delay, the officers granted approval and briefed their colleagues about the study. We were then directed to staff members who would complete the questionnaires. The staff also encouraged patients to participate in the study.

The healthcare professionals’ questionnaire was in English, while the patient questionnaire was translated into Chichewa and included both languages. Healthcare professionals completed their questionnaires independently during our site visit, and we collected them immediately or by day’s end. Many professionals praised the study’s importance and provided additional comments. Patients, assisted by a data collector, completed their brief, two-page questionnaires in the waiting room or after consultations, before or while waiting for their medications.

We maintained a diary with notes on each health facility visited, detailing their catchment areas, busyness, and whether they offered free or paid services. We also recorded the types of patients, such as mothers with babies. The diary included information on the paperwork required for study permission, the level of engagement with staff and patients, and any questions, comments, and observations about the questionnaire content.

### 3.3. The construction of the questionnaires

For each respondent, health professionals or patient, we collected demographic data on age, gender, home village, occupation, years in occupation, and highest qualification, but we did not record names. Participants were asked to specify their mother tongue and the language they used most frequently for communication.

We asked several questions about the use of language for learning and communication for healthcare professionals: in what language are lectures conducted and notes provided. Additionally, we inquired about language switching between English and local languages. We also inquired about the language of used in work communication for official reports, in trainings and seminars, and language used for communications among colleagues and with patients either verbally or via the patients’ health passports.

We conducted a vocabulary check for both professionals and patients. Professionals provided the Chichewa equivalents for selected medical terms related to diseases, medical tests, conditions, and symptoms, and rated the difficulty of translating these terms. The terms included abscess, biopsy, edema, fracture, hypertension, outpatient, embolism, seizures, gland, fever, and runny nose, all common in Malawi. Pulmonary embolism, a known COVID-19 complication, has high in-hospital mortality [16], [17] and is often associated with HIV and cancer. Edema, or swelling, is linked to conditions like pregnancy and diabetes, and can affect the eye. Biopsies, used in testing for diseases including cancer and skin conditions, are invasive and sensitive procedures. Misdiagnosis is a significant issue in Malawi, making early biopsy recommendations crucial [18], as discussions about biopsies can be lifesaving.

We provided patients with two lists containing five terms each, one for external organs and one for internal organs. We asked them to rank the difficulty of translating terms for organs and symptoms into Chichewa and to provide the equivalent translations. We inquired about their knowledge of the symptoms characterizing their conditions and about their ability to describe and discuss these symptoms in English, Chichewa, or other local languages, including whether they knew direct Chichewa equivalent terms. We also asked if language created challenges in communicating symptoms to clinicians and whether they understood what was written in their health passports.

The final three questions for professionals inquired about the opinion of healthcare professionals on the usefulness of a medical vocabulary to support them in translating or communicating in Chichewa and English.

### 3.4. Data Analysis Methods

We entered and analysed the data using Excel, applying codes and standardizing values into core categories. Occupations for healthcare professionals were categorized by role and seniority. For instance, Public Health Officers were grouped as Environmental Officers or Disease Control Assistants, and Medical Assistants were grouped as Health Assistants. We categorized occupations under the most relevant name, such as classifying a respondent stating “Biomedical Sciences” under Laboratory Technician. Seniority designations, like Senior Clinical Officer, were categorized simply as Clinician. Microsoft Excel 2016 was used to calculate the frequency of responses and present the data in tables. For analyzing Chichewa translations, we used clustering to merge similar terms and correct spelling mistakes. For example, “kutentha thupi,” “tentha thupi,” and “kutentha kwa thupi” were all recognized as “fever,” with the latter being more grammatically correct.

All responses given by participants in Chichewa were translated into English for frequency analysis. At question 5, patients were asked to select one of more diseases from a list and to give in Chichewa the symptoms that they knew were associated with those conditions. The responses were recorded in Chichewa, and we translated them into English, then we grouped and counted frequencies. The translation into English was based on meaning. For example, the following Chichewa terms mean “diarrhoea”: *kutsekula m’mimba, chimbuzi pafupipafupi, kupitapita ku chimbuzi, kupita kuchimbuzi mowirikiza, chimbuzi cha madzi madzi*. The literal translation of the first expression is “opening of the bowels/stomach”, and the next three refer to frequent visits to the toilet, while the last talks about watery stools.

### 3.5. Quality of the data

The credibility of our findings was increased by gathering information from several health facilities and gathering perspectives from both healthcare professionals and patients. We ensured our findings could be reliably applied and verified in other contexts by thoroughly describing the study environment and clearly outlining our methods, thus establishing a replicable audit trail for future researchers.

## 4. Results

We collected data from 20 clinics in Zomba district and 2 clinics in Machinga at the border with the Zomba district. In total we obtained responses from 79 professionals and 312 patients.

### 4.1. Demographic data for healthcare professionals

Table 1, Table 2, Table 3 and Table 4 contain the results from health professionals. Out of 79 medical professionals, 30 were female (representing 38% of the total) and 49 were male. Most were educated up to the diploma level (50), with a few having a degree(10) and 19 were educated at MCSE or certificate level. The clinics in the urban areas of Zomba City and Domasi, had the largest staff with degrees (9 out of a total of 10 participants with a degree).

**Table 1.** Demographics for Healthcare professionals. Average literacy levels of patients were estimated by healthcare professionals at each clinic.

| Question number |  | 1.2 | 5.1 | Highest Qualification (1.7) |  |  | Occupation (1.4) |  |  |  |  |  |  |
| --- | --- | --- | --- | --- | --- | --- | --- | --- | --- | --- | --- | --- | --- |
| Clusters and Clinics | Responde | Female, N | Avg. Literacy (5.1) | Degree | Diploma | Certificate or MCSE | Clinician | Laboratory Technician | Medical Assistant | Nurse | Nurse/ Midwife | Pharmacist | PHO |
| **Chingale** | **11** | **4** | **66%** |  | **7** | **4** | **3** |  | **1** | **4** | **3** |  |  |
| CHIPINI | 4 | 1 | 68% |  | 2 | 2 | 2 |  |  |  | 2 |  |  |
| MMAMBO | 5 | 2 | 65% |  | 4 | 1 | 1 |  |  | 4 |  |  |  |
| NKASALA | 2 | 1 | 65% |  | 1 | 1 |  |  | 1 |  | 1 |  |  |
| **City** | **38** | **13** | **66%** | **6** | **25** | **7** | **12** |  | **3** | **8** | **8** |  | **7** |
| CHANCO | 5 | 3 | 96% | 2 | 3 |  | 2 |  |  | 1 | 2 |  |  |
| CHULUCHOSEMA | 2 |  | 30% |  | 2 |  |  |  |  | 1 | 1 |  |  |
| COBBE | 4 |  | 60% |  | 4 |  | 2 |  |  | 1 | 1 |  |  |
| MATAWALE | 5 | 4 | 72% | 1 | 3 | 1 | 1 |  |  | 2 | 2 |  |  |
| NAISI | 3 | 1 | 63% | 2 |  | 1 | 1 |  | 1 | 1 |  |  |  |
| NAMADIDI | 2 | 1 | 57% |  | 1 | 1 | 1 |  |  |  | 1 |  |  |
| SADZI | 4 | 1 | 70% |  | 2 | 1 | 1 |  | 1 |  |  |  | 2 |
| ZILINDO | 3 | 1 | 60% |  | 2 | 2 | 1 |  | 1 | 1 |  |  |  |
| POLICE HOSPITAL | 5 | 1 | 65% | 1 | 4 |  | 2 |  |  | 1 | 1 |  | 1 |
| CITY CLINIC | 5 | 3 | 57% |  | 4 | 1 | 1 |  |  |  |  |  | 4 |
| **Domasi** | **25** | **10** | **41%** | **3** | **15** | **7** | **10** | **2** | **2** | **5** | **4** | **1** |  |
| BIMBI | 4 | 1 | 25% |  | 4 |  | 1 | 1 |  | 1 | 1 |  |  |
| HAWARD PAKER | 3 | 2 | 58% |  | 2 | 1 | 1 |  |  | 2 |  |  |  |
| MACHINJIRI | 5 | 3 | 50% |  | 3 | 2 | 1 |  | 1 |  | 1 | 1 |  |
| NAMASALIMA | 4 | 1 | 32% |  | 1 | 3 | 2 |  | 1 |  | 1 |  |  |
| DOMASI (GENES) | 4 | 2 | 42% | 2 | 1 | 1 | 2 |  |  | 1 | 1 |  |  |
| ST LUKES HOSPITAL | 5 | 1 | 41% | 1 | 4 |  | 3 | 1 |  | 1 |  |  |  |
| **Likangala** | **1** |  | **50%** |  | **1** |  |  |  | **1** |  |  |  |  |
| CHAMBA | 1 |  | 50% |  | 1 |  |  |  | 1 |  |  |  |  |
| **Machinga** | **4** | **1** | **46%** | **1** | **2** | **1** | **2** |  | **1** | **1** |  |  |  |
| MACHINGA HEALTH CLINIC | 2 | 1 | 35% |  | 1 | 1 | 1 |  |  | 1 |  |  |  |
| GAWANANI HEALTH CENTER | 2 |  | 57% | 1 | 1 |  | 1 |  | 1 |  |  |  |  |
| **Total** | **79** | **30** | **57%** | **10** | **50** | **19** | **27** | **2** | **8** | **18** | **15** | **1** | **7** |

**Table 2.**
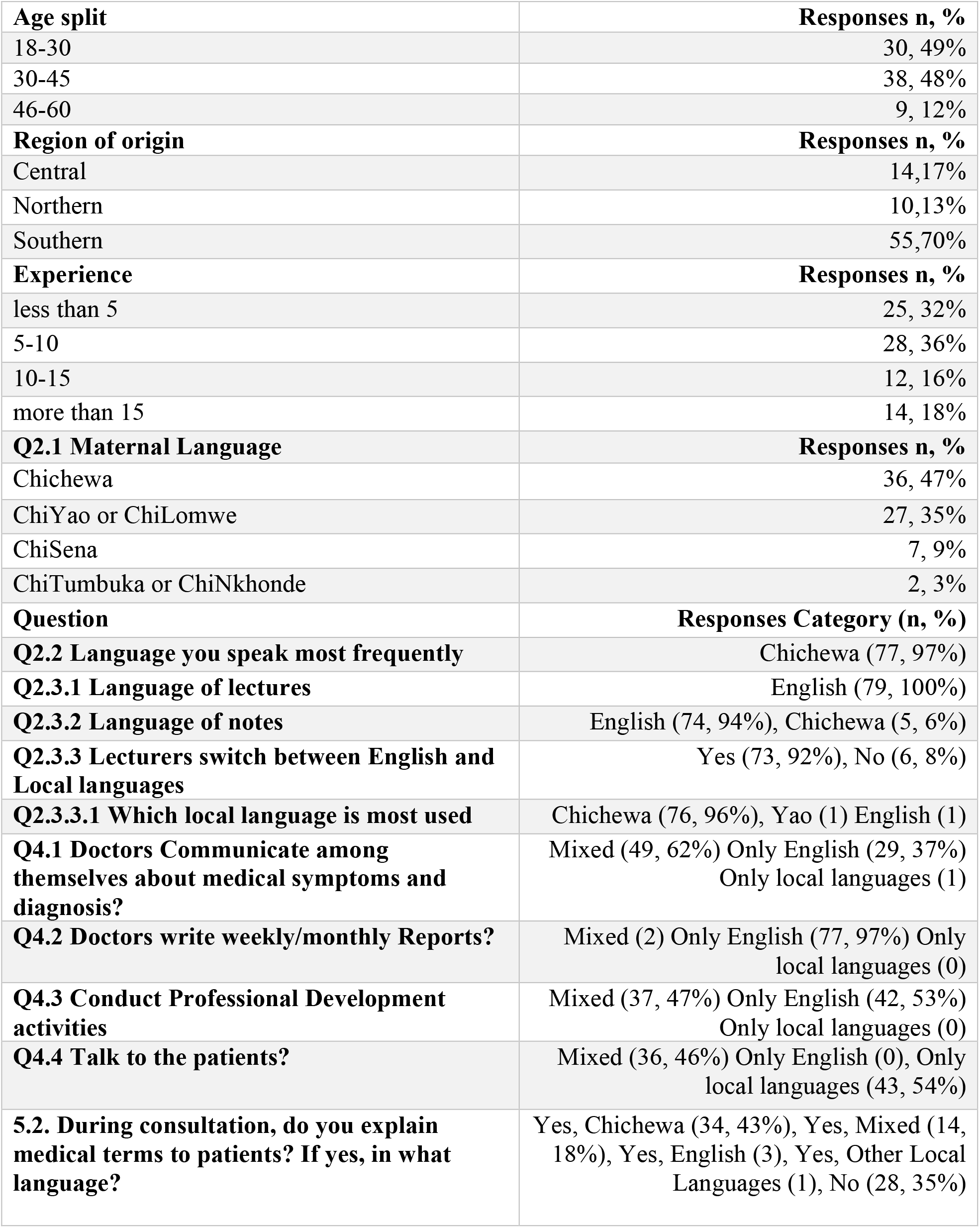
Profile of healthcare professionals. (C: Chichewa, E: English, Y: chiYao, L: chiLomwe)

**Table 3.** Which of the following affect your choice of language when talking to a patient? (rank from 1 to 3 = High impact) (Q5.4) In what language do you write in the health passport of the patient (Q6.1); What language do you use to record your own notes about the patient’s condition? (Q6.2) Do you use the notes from previously recorded in patients’ health passports (Q6.3), Do patients understand what is written in the health passport (Q6.4), Are patients open enough to disclose their symptoms to the clinician? (Q6.5) In a situation of critical illness, where the patient doesn’t not or cannot speak, what will you, as a clinician, rely on, in terms of symptoms and diagnosis? (Q6.6) Do you have a medical dictionary (Q7) How often do you use it (Q8) How useful do you consider a medical dictionary of English and Chichewa to be (Q9)

| **Q5.4.** | **Avg. Rank** |
| --- | --- |
| Nature of Sickness | 1.68 |
| Patients' level of literacy | 2.55 |
| Challenges in finding explanations in local language | 2.50 |
| Challenges in finding medical terms in local language | 2.71 |
| Time constraints | 1.58 |
| **Q6.1 Language on notes in health passport** | **Responses, n, %** |
| English | 78, 99% |
| Local Languages | 1, 1% |
| **Q6.2 Language for own notes about patients** | **Responses, n, %** |
| English | 79, 100% |
| Local Languages | 0, 0% |
| **Q6.3** | **Responses, n, %** |
| Sometimes | 34, 43% |
| Always or often | 43, 54% |
| Very rarely | 2, 3% |
| **Q6.4** | **Responses, n, %** |
| Yes, largely | 16, 20% |
| Very little | 57, 72% |
| No | 6, 8% |
| **Q6.5** | **Responses, n,%** |
| Yes | 29, 37% |
| Most | 5, 6% |
| Mostly women | 13, 16% |
| Mostly women and the elderly | 4, 5% |
| Depends on literacy, language, education, age, gender | 27, 34% |
| No | 1, 1% |
| **Q6.6** | **Responses, n, %** |
| The patient's facial expressions and gestures? | 25, 32% |
| Physical examination of the patient | 58, 73% |
| What the guardian will narrate to you based on what the patient confided to that guardian? | 31, 39% |
| Where available you rely on lab results | 42, 53% |
| **Q7. Do you have a medical dictionary Q8. If yes how often do you use it?** | **Responses, n, %** |
| No | 51, 65% |
| Yes, frequent use | 6, 8% |
| Yes, rarely use it | 8, 10% |
| Not asked this question | 14, 18% |
| **Q9. Usefulness of a medical dictionary** | **Responses, n,%** |
| Positive | 61, 77% |
| Negative | 1, 1% |
| No response | 3, 4% |
| Not asked this question | 14, 18% |

**Table 4.**
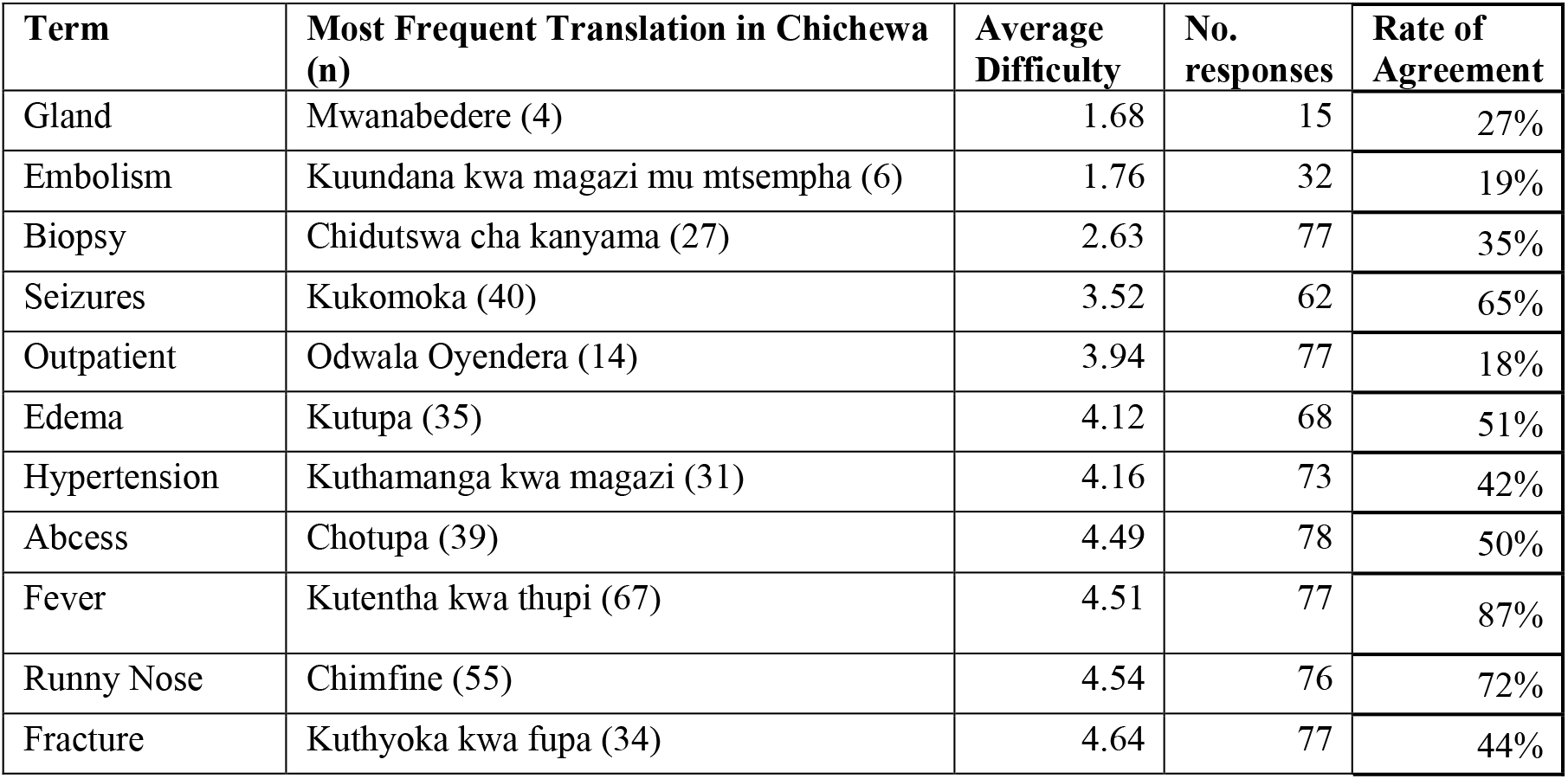
Medical terms translations to Chichewa. The table is sorted in terms of average difficulty, (ordered from highest to lowest). The difficulty scale used was 1= Very Difficult and 5 = Very easy.

In the more rural clinics (excluding Machinga), there were no staff trained at the degree level. The most common occupation was Nurse (33), who perform dual nursing/midwifery roles in Malawi. The second-largest category was Clinicians (27), which included clinicians, clinical officers, and the more senior clinical technicians. Public Health Officers (7) and Medical Assistants (8) followed. Two respondents from Domasi were Laboratory Technicians, and there was only one pharmacist. These occupations reflect the availability of staff during our visits and reasonably represent the services at typical rural clinics in Malawi. Most rural clinics lack laboratory testing facilities, but St. Luke’s Hospital in Domasi, a larger site, offers paid laboratory services. Additionally, most rural clinics operate without a pharmacist, a role requiring a degree, but may have assistant pharmacists[19].

Almost half (49%) of the respondents were between 30-45 years old, 12% were ages between 45 and 60 and 39% were ages between 18-30. This covered a good time span, indicating that our respondents experienced various types of training over approximatively 20 years. They also had a good mix of work experience: 26 out of 79 (33%) had more than 10 years of experience in their role and 32% has less than 5 years of experience.

Most healthcare professionals came from the Southern region (69%), a third came from the Central and Northern regions (31%). This implies an interesting mix of languages, dialects, and cultural norms. Table 2 shows that less than half of the respondents, 47% (36 out of 79) had Chichewa or its dialects as their maternal language. More than a third (34, or 35%) had Chiyao, Chilomwe or Chisena as their mother tongues. For the rest (9), Chitumbuka, Chitonga and Chinkhonde (the languages of the Northern region) were the mother tongues. Most participants were from the Southern Region and 30% had moved from the Central or the Northern region to live and work in the South. This reflects a large mobility of people as their pursue careers and jobs across Malawi. Regardless of their mother tongue, for most of the respondents (97%), Chichewa was the language they spoke most frequently. This reflects the role of Chichewa as the lingua franca in Malawi [20].

#### Teaching and learning for healthcare professionals are done solely in English

As shown in Table 2, English is the language of teaching/instruction. All respondents indicated that lecturers are done in English and 95% responded that notes are also provided in English. Lecturers frequently switch between English and Chichewa to provide additional explanations to their students. Majority (97%) indicated that the language they use the most for communication is Chichewa (responses to Q2.2). Unlike lectures, training is conducted in mixed English and local languages (mainly in Chichewa). It was not clear whether the content expressed in English and that expressed in Chichewa overlap, but what is evident from the responses is that Chichewa is better for facilitation and for creating a relaxing and more approachable atmosphere during workshops. The following were mentioned by our respondents as factors for the use of mixed English and local languages during training:

> “To rouse the interest of the workshop participants / provoke the interest of learning process.” (Respondent 2, Machinjiri).
>
> “When expressing a practical scenario as an example on the ground in the process of training or imparting knowledge to each other during professional development activities such as workshops.” (Respondent 4, Namasalima)
>
> “For easy communication in sharing experiences on symptoms and diagnosis. To clarify a point that has a practical example in experiences obtained.” (Respondent 3, Naisi)
>
> “When they want to emphasise a point and make sure it is well understood. When addressing workers of different cadres, especially the lower cadres.” (Respondent 9, Domasi Genes)

#### Communication in the workplace among healthcare professionals when discussing patient conditions and medical history takes place in English with mixed use of local languages

English took a central role when doctors communicated among themselves about medical symptoms and diagnosis: 62% participants used mixed English and Chichewa and 37% responded that they used only English (Q4.1). Only one participant indicated the use of only local languages, possibly caused by a misreading of our question. Healthcare professionals preferred the use of English because some medical terms were very difficult to find in Chichewa and did not align well with equivalent concepts in local languages for example “intrapartum haemorrhage”. Chichewa was also used alongside English for easy understanding with each other on medical issues for example “stethoscope *iyi siikugwira ntchito ka clot ka magazi kanatseka mtsempha wa magazi ku ubongo*” meaning that the “stethoscope would not work or make a sound when patients may have a clot that blocks the blood flow to the brain”. (Respondent 3, Chipini)

#### Communication with patients requires the use of the patient’s language, which may be different than that of the healthcare professional

When communicating with patients, it was seen as imperative to be able to use the language of the patient or that of the guardian to explain medical conditions or treatment administration. But some professionals confessed that they sometimes use English intentionally to hide painful or sensitive information from the client about their condition.

> *“In circumstances where the term would best be described in Chichewa, Chichewa is used in mixture with English e*.*g. explaining some routes of drug administration like vaginal and anal*.*” (R1, Machinjiri)*
>
> *“I work here at the health centre, let us assume in an emergency case that I need to explain to the patient and guardian the condition plan of care and reason for a referral*.*” (R2, Bimbi)*
>
> *“Patients are also of different academic background that determines their understanding and way of communicating to them. It is also very difficult to communicate to patients with special needs*.*” (R3, Police)*
>
> *“It depends on the literacy level of that patient and while observing the patient’s privacy and in respect to the language of that particular patient*.*” (R2, Chipini)*

Communication between doctors and patients took place predominately in the local languages (54% of respondents) and mixed English and local languages (36% of the respondents) and no one indicated the exclusive use of English (Q4.4).

#### Medical or hospital notes and reports are written only in English. Notes, diagnoses, and medication recorded in the patients’ health passports are in English

English was the language of report writing: 97% indicated that they used only English when writing reports (Q4.3). English was also the sole language for making entries in the patient’s health book (Q6.1): 78 participants (99%) responded that they wrote notes in the health passports in English and only one participant responded that he/she used local languages, but this could have been recorded erroneously. English was unanimously the language in which healthcare professionals wrote their own personal notes about the condition of their patients (Q6.2).

#### Patients rarely know what is written in their health passport

A health passport book is a paper-based, patient-kept portable medical record. Patients carry their health book with them when they visit a healthcare provider, who in turn write details of the visit in the passport. Malawi uses health passport books in all its public facilities. Sample copies of pages in health passports are given in **Error! Reference source not found**. and **Error! Reference source not found**..

In the opinion of healthcare professionals, patients (Q6.4), *“as many did not go to school hence many cannot understand. Doctors use symbols and signs that can be well understood by health professionals and not everybody else”. “The entries made in the health books are mainly meant for the health professionals so rarely can that be understood by the patient who in majority not familiar with signs and symbols used. But the patient has a duty to remember the descriptions made on symptoms he/she confided to that clinician during consultation*.*” “Some patients come for consultation without a health book. In this case, a clinician relies on physical examinations, lab results and probing the past health history which results in errors and exaggerations. Probing is possible only for a patient who is not critically ill and not unconscious or through the guardian, the usage of a guardian is also subject to exaggerations”* (C1, Howard Parker).

#### Healthcare professionals use notes written in previous visits in the health passport of the patient

Almost half (46%) of healthcare professionals said that they rarely or sometimes use the previous notes written in the health passports and 54% said that they often or always referred to notes written in previous visits (Q6.3). Given the importance of the health passports as containing the most complete medical history of the patient, we expected this number to be higher. For the great majority of patients in Malawi, no case notes or record cards are kept on the premises by hospitals, health centres and other facilities. Our findings may be explained by the nature of the visits or notes that were made or simply by the fact that there is a loss of confidence in the notes recorded in these health passports and that they tend to be incomplete [21].

#### Healthcare professionals do not often explain medical terms to their patients

Openness towards healthcare professionals was influenced by literacy, language, education, age, and gender of the patients. The women and the elderly were said to be more comfortable in expressing their symptoms (Q6.5). When asked whether they explain to their patients’ medical terms relevant for their condition and diagnosis, 63% of the healthcare professionals answered that they do that under some conditions and they use mainly Chichewa or English (Q5.2 and Q5.3). However, they had challenged in expressing medical terms or finding explanations for diseases or conditions in local languages and this affected their ability to use local languages (Q5.4).

> *“Yes, however I don’t prioritise on giving explanations on medical terms because majority of the patients will not understand many technical medical terms though sometimes, I explain the diagnosis to some patients just for prescription*.*” (C1, Howard Paker)*
>
> *“No, some medical terms remain so technically scientific that they do not have direct and precise equivalent terms in local languages such as in Chichewa or Lomwe. Description of medical terms are rarely done; most doctors prioritise telling the patient the lab results and the way the medicine given be taken at home*.*” (C2, Howard Packer)*
>
> *“Not terms but we tell the patient the diagnosis either in Chichewa or the language of that particular patient*.*” (C3, Zilindo)*
>
> *“Yes, depending on the literacy level of a particular patient because to an illiterate, it will appear a mockery*.*” (C1, Mmambo)*

#### The impact of patients’ literacy

Patients’ literacy affected the extent of explanations that healthcare professionals gave to their patients, and it influenced also the relevance to the patient of notes written in the health passport (34% indicated this on Q6.5). We asked our respondents to estimate the level of literacy of their patients. Very few sites had a literacy rate of over 70%. The Chanco clinic was an exception because it served the University of Malawi staff and students. The literacy rate averaged 60% in Zomba City and Chingale cluster (this is situated near the Zomba city), and around 50% in more rural clusters.

Because literacy is a key determinant of health, we can anticipate that the declining literacy leads to declining health outcomes. Official statistics indicate that Malawi’s adult literacy rate, defined as the “percentage of people 15 and above who can both read and write, understanding a short simple statement about everyday life” was 64 percent in 1998 and declined to 62 percent by 2015, the most recent year for which figures are available^1^. This is marginally better than similarly situated Mozambique (60%) but not nearly as high as Tanzania (77%) or Zambia (87%). It is possible that literacy as defined by official statistics does not reflect health literacy. A small study in Malawi showed that generally in Malawi patients lack access to health information in a language and format that they can understand [22] and that among young adults who completed the final year of primary school (grade 8), only 40 percent could fully read and comprehend Chichewa and that just over one-half of the young adults demonstrated full reading and comprehension. In Zambia an important segment of the literate population, especially those who did not proceed past the primary school level, preferred to read about health matters in their mother tongue [23]. We can conjecture that this is also the case in Malawi.

#### Healthcare professionals do not have access to medical dictionaries

Most respondents (65%) indicated that they did not have access to a medical dictionary (Q9). Among those who said that they do have a dictionary, only 6 respondents said that they use it. The majority (77%) of respondents agreed with the importance of having access to a bilingual English-Chichewa medical vocabulary for their work:

> *“A medical dictionary in a mobile phone or online is very useful because it is not all the time that you remember the doses of drugs especially in children, so the dictionary helps and also in times where the condition is new to the area, we consult the dictionary*.*” (C1, Machinjiri)*
>
> *“It would be useful because sometimes you may come across a rare condition that needs dictionary consultation*.*” (C2, Bimbi)*
>
> *“It would be very useful. It will make it easy to explain some of the medical terms which lack direct translations*.*” (C2, Chanco)*
>
> *“It will help in making work easier and promote understanding and cooperation between health workers and patients*.*” (C2, Mmambo)*
>
> The respondent who answered negatively thought that such a dictionary would be rarely used for common diseases which should be well known to healthcare practitioners, but that in cases of rare conditions such a dictionary would be useful.

> *“Very rarely will it be used. A clinician is supposed to know these symptoms by heart in readiness for unusual health cases. Of course, we will refer to it only in cases where the conditions are new, not familiar with and unique in nature. Frequent dictionary consultation will not help in emergency cases hence might delay the treatment*.*” (C1, Howard Paker)*

#### Healthcare professionals struggle to express medical terminology in local languages

We asked healthcare professionals to rate the difficulty and give the Chichewa equivalent for 10 medical terms. These referred to common conditions (embolism, abscess, fracture, hypertension), symptoms (fever, runny nose, edema), procedure (biopsy), anatomical part (gland) and hospital related term (outpatient). The list was kept short so that the respondents would be able to fill it in during their work break. Responded commented positively on the choice of terms on the list. Terms that were difficult to translate, received a low difficulty score. For example, the word *gland* was the most difficult to translate receiving a difficulty score of 1.68. Only 15 participants attempted to translate it and there was a high disagreement in terms of the translations given. This means that four people translated the word glad as “*mwanabedere*” in Chichewa and 11 other different terms were given.

Even terms that were easy to translate into Chichewa, such as *fever* or *runny nose*, received multiple translations. The most given Chichewa terms (67 mentions) for *fever* was “kuthentha kwa thupi”, but 11 other translations were given. 31 respondents agreed with “kuthamanga kwa magazi” as the translation for *hypertension* but 42 other responses were given. Hence, the rate of agreement in translating fever was 87% but the rate of agreement for translating hypertension was 42%. This means that fever was easier to translate to a direct Chichewa term than was hypertension. The rate of agreement was calculated by dividing the frequency of the most common translation to the total responses. For example, the agreement rate for hypertension was calculated as 31/73= 42%. The hardest terms to translate were *gland* and *embolism*. Only 4 participants managed to give the Chichewa equivalent for *gland* and only 6 managed to translate *embolism*.

This short vocabulary check illustrates the fact that healthcare professionals did not find it easy to give equivalent translations of English terms into local languages. Judging by the multiple different translations that were given even for easier terms, we can deduce that a medical dictionary with well thought translations and expressions would prove useful for healthcare professionals, who use English in all medical notes but must switch to Chichewa when speaking to patients.

### 4.2. Results from patients’ questionnaires

Table 5, Table 6 Table 7 contain the results from patients. We collected responses from 312 patients, 87 were men and 224 were women (representing 72% of the total). The gender split reflected the fact that many clinics prioritises maternal health and under 5 consultations. Half of the respondents were between 18-30 years (160), 36% were between 30 and 45 (113), 10% were between 46-60 years of age (30) and 3% were above 60 (9). The top 10 occupations were farming (93), housewife (70), business (63), student or pupil (41), teacher (12), piecework (9) and house maids (3) and fishing and forestry (4).

**Table 5.** Patients demographics.

| Facility | Patients, n | Gender |  |  | Education |  |  |  |  |  |  |
| --- | --- | --- | --- | --- | --- | --- | --- | --- | --- | --- | --- |
|  |  | Female, n | Male, n | Unknown, n | Not attended | <PSLC | PSLC | JCE | MSCE | Certificate | Diploma |
| **Chingale** | **56** | **53** | **3** |  | **6** | **11** | **16** | **12** | **11** |  |  |
| Changalume | 16 | 15 | 1 |  | 1 | 2 | 4 | 4 | 5 |  |  |
| Chipini | 7 | 6 | 1 |  |  |  | 4 | 2 | 1 |  |  |
| Mmambo | 15 | 15 |  |  | 1 | 5 | 4 | 2 | 3 |  |  |
| Nkasala | 18 | 17 | 1 |  | 4 | 4 | 4 | 4 | 2 |  |  |
| **City** | **167** | **108** | **59** |  | **19** | **8** | **57** | **37** | **43** | **2** | **1** |
| Chanco | 12 | 3 | 9 |  |  |  |  |  | 12 |  |  |
| Chuluchosema | 15 | 11 | 4 |  | 1 | 2 | 7 | 4 | 1 |  |  |
| City Clinic | 15 | 8 | 7 |  | 2 |  | 4 | 5 | 4 |  |  |
| Cobbe | 12 | 8 | 4 |  |  |  | 3 | 5 | 4 |  |  |
| Matawale | 20 | 17 | 3 |  |  |  | 10 | 3 | 6 |  | 1 |
| Naisi | 17 | 10 | 7 |  | 2 |  | 5 | 4 | 6 |  |  |
| Namadidi | 19 | 10 | 9 |  | 4 |  | 8 | 2 | 3 | 2 |  |
| Police Hospital | 15 | 11 | 4 |  | 2 |  | 6 | 5 | 2 |  |  |
| Sadzi | 19 | 14 | 5 |  | 1 | 3 | 6 | 5 | 4 |  |  |
| Zilindo | 23 | 16 | 7 |  | 7 | 3 | 8 | 4 | 1 |  |  |
| **Domasi** | **79** | **56** | **22** | **1** | **19** | **20** | **15** | **11** | **12** | **1** | **1** |
| Bimbi | 20 | 15 | 5 |  | 4 | 3 | 6 | 5 | 1 | 1 |  |
| Domasi (Genes) | 3 | 2 | 1 |  |  | 1 | 1 | 1 |  |  |  |
| Haward Parker | 3 | 3 |  |  |  | 1 | 1 |  |  |  | 1 |
| Machinjiri | 25 | 16 | 9 |  | 11 | 7 | 2 | 2 | 3 |  |  |
| Namasalima | 22 | 16 | 6 |  | 4 | 6 | 3 | 3 | 6 |  |  |
| St. Lukes Hospital | 6 | 4 | 1 | 1 |  | 2 | 2 |  | 2 |  |  |
| **Likangala** | **9** | **7** | **2** |  |  | **6** | **1** | **2** |  |  |  |
| Chamba | 9 | 7 | 2 |  |  | 6 | 1 | 2 |  |  |  |
| **Machinga** | **1** |  | **1** |  |  |  | **1** |  |  |  |  |
| Machinga Health Clinic | 1 |  | 1 |  |  |  | 1 |  |  |  |  |
| **Total** | **312** | **224** | **87** | **1** | **44** | **45** | **90** | **62** | **66** | **3** | **2** |

**Table 6.** Knowledge of conditions and symptoms. 5i = Can you describe well the conditions affecting you? 5ii = How far do you know the symptoms of the conditions you suffer from? 5iv = Do you use direct Chichewa concept equivalent to English in expressing symptoms? 6i = Do you find it easy to express your symptoms to the clinician? 6ii = Does language creates a challenge in expressing these symptoms? 6 iv = Do you understand what the doctor writes in your health book? For questions 5 ii – 6 iv 19 respondents were not asked these questions.

| **Q5.i. (can describe symptoms and conditions)** | **Respondents, n** |
| --- | --- |
| No | 5 |
| Yes | 14 |
| Yes, in Chichewa | 260 |
| Yes, in English | 12 |
| Yes, in other language: Yao | 12 |
| Yes, in other language: not specified | 2 |
| Yes, in other language: Lomwe | 6 |
| Yes, in other language: Tumbuka | 1 |
| **Q 5ii (knows symptoms for conditions)** | **Respondents, n** |
| I know all of them | 10 |
| I know few of them | 152 |
| I know many of them | 131 |
| **Q 5 iv (use direct Chichewa terms for conditions or symptoms)** | **Respondents, n** |
| (blank) | 5 |
| No | 276 |
| Yes | 12 |
| **Q 6i (finds easy to express symptoms)** | **Respondents, n** |
| Yes, always | 84 |
| Yes, sometimes | 167 |
| No | 42 |
| **Q 6ii (language is a challenge in expressing symptoms to doctors)** | **Respondents, n** |
| Yes | 185 |
| Yes, sometimes | 10 |
| No | 96 |
| (blank) | 2 |
| **Q 6 iv (understand doctor's notes in health passports)** | **Respondents, n** |
| No | 232 |
| Yes | 53 |
| Yes, a little | 4 |
| (blank) | 23 |
| **Grand Total** | **312** |

**Table 7.**
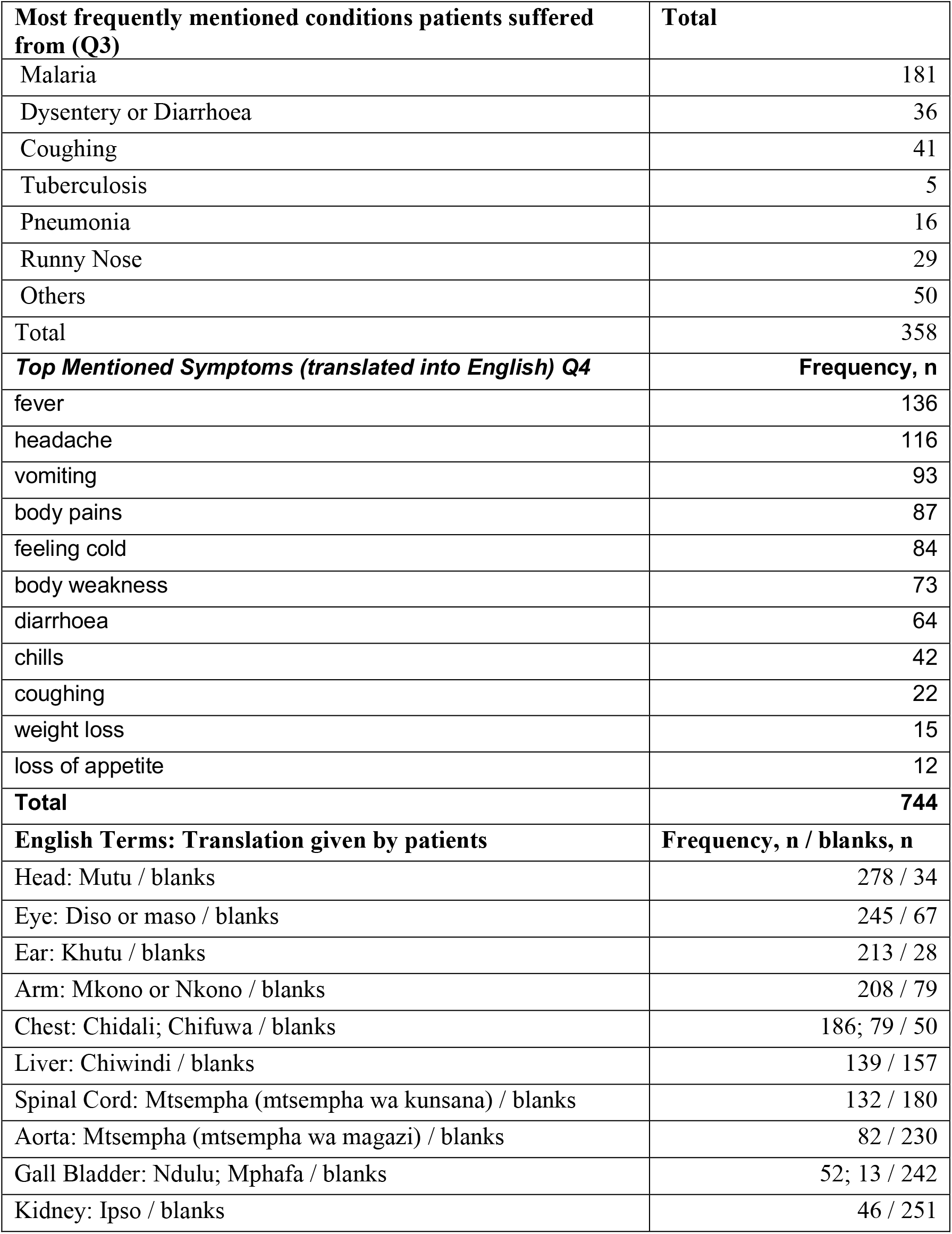
Most frequently mentioned symptoms.

More than half (57%) of all patients were educated only up to Primary School Level Certificate level (179), 20% were educated at JCE level. Some respondents had never been to school or have dropped out of school very early (44, 14%). Interestingly, in Zomba City and Domasi, both clusters with good access to schools, had the highest proportion, 11% and 24% of the respondents respectively, who did not attend school. Primary School Leaving Certificate of Education (PSLCE) examination are written by learners who complete their final grade at primary school. JCE is obtained after completing two years of secondary school, hence it is an intermediate certificate between PLSCE and MSCE.

Only 20% of patients obtained an MCSE certificate and only 2% were educated at diploma level. The Malawi School Certificate of Education (MSCE) examination is the final school examination and is written by students who are completing the fourth year of secondary school. Students who pass this examination qualify for selection into public and private colleges and universities or get absorbed into the job market. Both Chichewa and English are subjects for examination but only English requires a pass as a condition for the award of all education certificates.

#### Communication with the doctor / healthcare professionals: patients struggle to express medical terminology in local languages

Most patients (83%) can describe the symptoms and conditions they suffer from in Chichewa, but very few can do so comfortably in English (4%) (Q5.i). Half of the patients said that they knew a few of the symptoms that characterise the conditions they suffer from, and 42% said that they knew many of them (Q5ii). Most respondents answered that they do not use direct Chichewa terms conditions and symptoms (87%) but use descriptive language (Q5 iv).

At question 6i, we asked patients if they found easy to communicate and describe their symptoms to healthcare professionals. 29% responded that they always found it easy, 57% indicated that it was sometimes easy and 14% responded indicated they did not find it easy. Those who responded “Yes, sometimes”, mentioned several factors that affected their openness or ability to communicate: the nature of their disease, the rapport that was created and language differences between them and healthcare professionals. Those responding “No”, said that the doctors’ attitude was a major inhibitor of their openness: “*No, it’s difficult because of fear and shame and the doctor yelling*.*”* Here are some patients’ quotes highlighting these challenges:

> *The doctor might not understand me if I speak Lomwe while he instructs me in Chichewa. (P2, Machinjiri)*
>
> *When I speak Yao, the doctor won’t understand it then I force myself in difficulty to express it in Chichewa. (P7, Machinga)*
>
> *There may be a problem because they cannot understand each other, and the doctor may write the wrong diagnosis. (P31, Bimbi)*
>
> *The doctor is unfriendly, he appears very angry and unapproachable and shouts at patients, so patients do not have an opportunity to freely express their symptoms. (A dotolo amaoneka okwiya, ansunamo, opanda nsangala a mangawa ndipo amangokalipira potilankhula choncho sitimakhala ndi mpata onena matenda anthu momasuka*.)

Some indicated that there were also problems in “listening to each other” *(Simungamvane polankhulapo)* and in taking sufficient time to understand each other’s signs and gestures *(sitimvetsetsana zizindikiro)*.

*When a patient is expressing his/her symptoms he [the clinician] does not pay attention and appears not to be listening. (Pamene tikufotokoza matenda, samawonetsa chidwi pomvetsera zizindikiro zomwe tikunena ndipo amaoneka kuti sakukumvetsera*.)

> Some diseases were difficult to explain *(matenda ena ngovuta kuwafotokoza)* and patients failed to explain their illness due to a lack of appropriate words (*nthawi zina odwala malephera kzifotokoza matenda ake chifukwa cha kusowa mau oyenera kapena kudwalika)*.

*Privacy is compromised because the doctor’s desk is as a metre away from the next patient sitting on a bench on a queue. (*Sitimamasuka kuulula matenda chifukwa mpando ndi desiki ya adotolo yayandikirana ndi pamene odwala omwe ali pamzere akhala pa chi benchi, kudikira kuti nawonso alowe akumane ndi dotolo.)

Most patients (74%) did not know what was written in their health passport (Q 6 iv). They were able to occasionally recognise names of drugs and results of tests such as “Positive” or “Negative”. The reasons for this varied: sometimes this was caused by patients’ literacy levels, sometimes by “sloppy writing, the notes are illegible, and they (*the clinicians*) use medical symbols that I don’t know”.

> *I can’t understand it because I have not yet gone further with school (std 6). But also doctors write in English so fast and in an illegible joint letter. But if they were to make those entries in Chichewa, I would have asked a friend of upper class to read and interpret them for me. (P23, Machinjiri, translation)*
>
> *Not all doctors use medical terms, they write unreadable, they use symbols that I don’t know like RX, RCT, C/O, HPC, PDH. (P143, Police Hospital, translation)*
>
> *The doctor does not refer to or take the initiative to closely follow my health history as written in my health book. He does not open my health book back entered pages to check previous prescriptions but rather just opens a new page and makes new illegible scribbles*. (A dotolo samakhalanso ndi chidwi kuti aone matsamba ena a buku langa lakuchipatala kuti adziwe za matenda omwe ndinadwalapo mmbuyomu, ndipo mmalo mwake amangotsekula tsamba latsopano nalemba matenda a tsiku ili komano mosawerengeka.)
>
> *The doctor does not explain neither say to the patient what disease the patient is suffering from nor what (drug name) drugs has he prescribed in the health book. (*A dotolo samakuuza iwe odwala matenda omwe akupeza nawo komanso kukufotokozera mtundu wa mankhwala omwe akulembera kuti ukalandire pa windo.)
>
> This agreed with the responses from healthcare professionals, the majority (80%) responded that patients did not or understood very little what was written in their health passports (Q6.4). Moreover, 46% professionals said that rarely or only sometimes referred to notes in health passports (Q6.3).

#### Health Vocabulary

We asked patients to name in English ten health terms (Q3 and Q4). Their responses are summarised in Table 5 and were as follows: 278 patients translated the word head as *mutu* and there were 34 blanks (no answers); 245 people translated eye as *maso/diso* and there were 67 blanks. Chest was translated as *chidali by* 186 respondents, as *chifuwa* by 79 and there were 50 blanks. The table contains only the two topmost frequent Chichewa translations given by participants. The number of blanks is an indicator of the difficulty or unfamiliarity with either the English terms or the Chichewa equivalent. Surprisingly, many did know how to translate the word *kidney* and asked the researcher to demonstrate the position of the kidney in the body, to help them to identify it and try to find its corresponding Chichewa name. Liver is commonly seen in chickens and was easily recognised and translated to Chichewa. Overall, participants were familiar with the Chichewa and English terms for the most common external body parts but faced difficulties in naming internal organs.

#### Symptoms and conditions mentioned as being most familiar to patients

Most frequently mentioned as common diseases by patients were *malaria* followed by *coughing* and *dysentery or diarrhoea*. Under Other, HIV/AIDS (22) and BP (6) were the most mentioned conditions (Q5). The City clinic had the highest numbers of reported HIV/AIDS (14), followed by Chingale (8). This option was not on the questionnaire because we considered it to be sensitive.

However, several patients felt free to disclose that HIV was one they suffer from and were more familiar with its symptoms.

Patients were asked to list the symptoms they are most familiar with (Q5). They described over 160 different types of symptoms in Chichewa (over 1000). Some expressions in Chichewa may be translated to the same symptom in English but describe different “feelings” or perceptions of what the patient experienced. For example, “coughing” was described using several expressions indicative of how frequent or persistent the cough: *kutsokomola kwa nthawi yayitali* (coughing for a long time, a symptoms usually associated with TB, the meaning being a persistent cough lasting for longer than two weeks), *kutsokomola mowirikiza* (repeated coughing but over a short period of time), *kutsokomola pafupipafupi* (frequent coughing) *and kuwirikiza kutsokomola* (direct translation being “double coughing” and the meaning being cough that is prompted by itchiness in the throat). There were several terms used for diarrhoea, describing the length of time or the presentation and type of the stool. Some symptoms were known to be side effects of medication, especially those linked to ARV drugs, and referred to appearance, skin conditions and hair damage or loss. These were highly descriptive and do not have an equivalent medical symptom.

**Figure 2.**
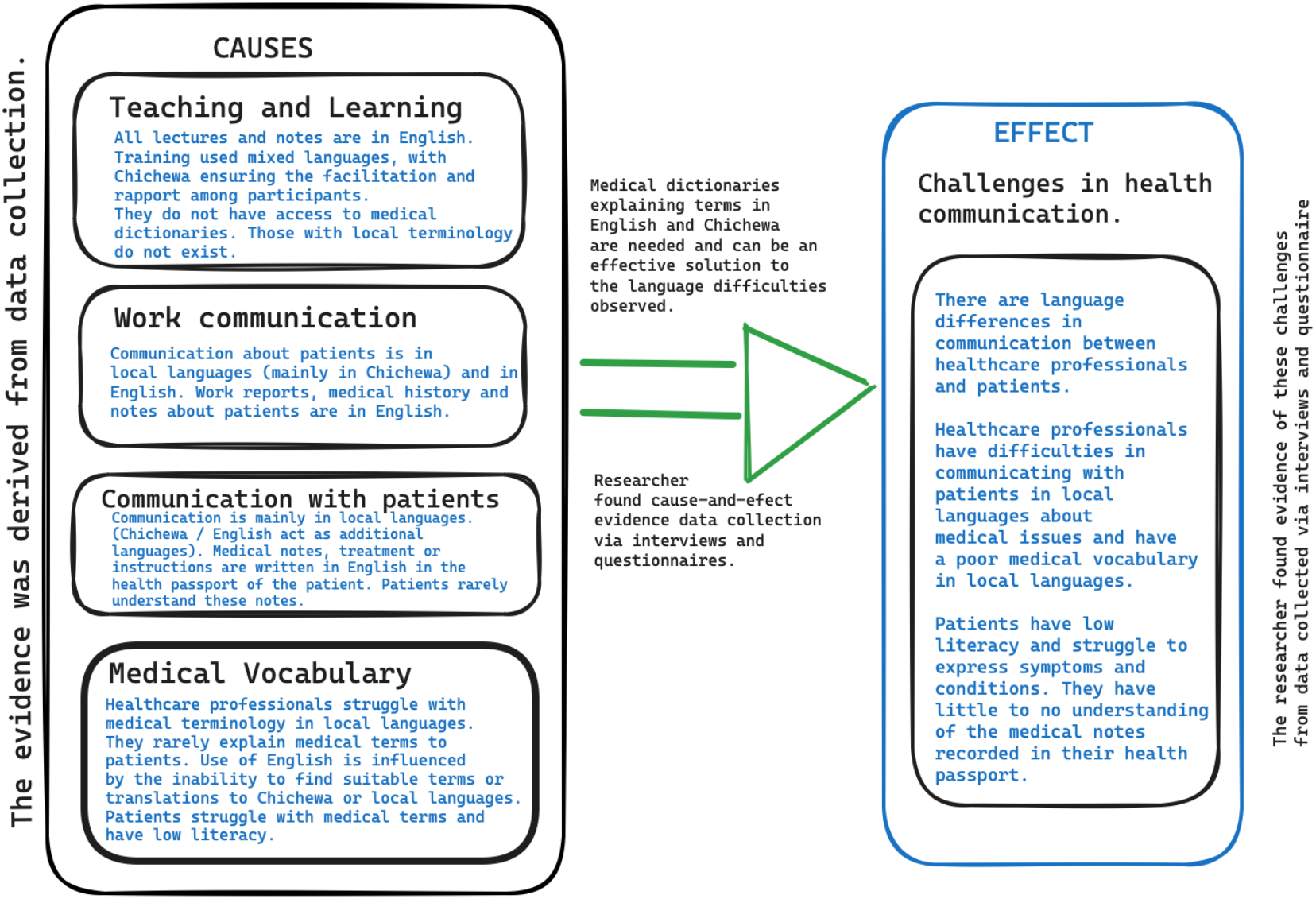
Cause-and-effect evidence from data collection in support of the hypothesis propositions.

## 5. Discussion

The results of this research form a strong cause-and-effect evidence to support the hypotheses that healthcare professionals are poorly prepared to communicate with patients in local languages and that language barriers occur from both the side of healthcare professionals and that of patients. These barriers have multiple causes coming from historical legacies, language policies, and social and educational structures. The role of English and that of local languages while largely unchanged in terms of policies and official use, has witnessed a shift over time in response to changes in social and educational structures. An observed decline in education and literacy leads to a much deteriorated access to knowledge. Revisiting language needs for health at facility level is possible, as out study demonstrated, and it is essential for ensuring equitable access and good quality of health.

The results of our investigation are summarised in Table 8 and show that English and local languages occupy different roles, often complementary and non-overlapping, in communication. English is the language of teaching and learning and for of official work communication. Communication about patients during work interactions between healthcare professionals relies on a mixed use of English and Chichewa. Chichewa and to a lesser extent other local languages is the main language used for communicating with patients verbally but written clinical history in health passports uses only English. The medical vocabulary is learned in English, thus finding Chichewa equivalent terms during consultations is challenging.

**Table 8.** Summary of results from questionnaire as they refer to the hypothesis propositions.

| Language used for tasks below (Responses, %) | English | Local Languages | Mixed |
| --- | --- | --- | --- |
| A. Teaching and learning for healthcare professionals. | 100% |  |  |
| C. Medical or hospital notes and reports. | 94% | 6% |  |
| E. Notes, diagnoses, and medication recorded in the patients' health passports. | 100% |  |  |
| B. Communication in the workplace among healthcare professionals when discussing patient conditions and medical history. | 37% | 1% | 62% |
| F. Communication with patients requires the use of the patient's language. |  | 54% | 36% |
| G. Healthcare professionals explain medical terms to their patients. | 4% | 43% | 18% |
| Responses, % | Yes | No |  |
| D. Healthcare professionals have access to medical dictionaries. | 22% | 78% |  |
| G. Healthcare professionals explain medical terms to their patients. | 65% | 35% |  |
| H. Healthcare professionals use notes written in previous visits in the health passport of the patient. | 54% | 46% |  |
| I. Patients know little or not at all what is written in their health passport. | 80% | 20% |  |
| J. Both healthcare professionals and patients struggle to express medical terminology in local languages* | 100% |  |  |

Our findings provide evidence for the need to establish better bridges between these languages. For example, nursing and medical colleges could introduce dedicated language courses that cover key competencies such as terms for organs of the human body, names of common diseases in vernacular; or language skills for medical interviewing, and patient-centred explanations of medical diagnoses, assessment and treatment plans. This would improve the appreciation that healthcare professionals of patients and lead to more listening and better two way communication during consultations.

Patients too struggle with language differences. Often, they speak a different mother tongue than the clinician. Falling literacy and education opportunities means that they vocabulary in local languages and in English is deteriorating. They have little to no understanding of what is written in their health passport. This is due to poor legibility of the writing but also due to the sole use of English for these notes. When the medical professionals take the time to explain their condition in more details, patients find it challenging to understand medical terms. As our research shows, the limited vocabulary in local languages of both the patient and of the healthcare professionals is a significant barrier. In addition, communication can be improved or deteriorated by the rapport that is being created during a visit. Most medical professionals take the view that it is enough to communicate the final diagnosis and treatment, rather than to explain in more details the medical aspects of the condition their patients suffer from. Some see this as off-putting for patients. However, in our pilot study we found that patients desire to gain better knowledge about diseases and other terms that affect their medical condition including knowledge about human organs and their function.

When healthcare professionals and patients meet during repeated visits as it is the case of patients enrolled in specific treatments that require follow-ups and checkups, e.g., HIV/ART treatments, there is a better chance to establish a common vocabulary [24]. In most cases, communications that take place during the first visits may be the only opportunity for dialog and are crucial for early diagnosis and for setting the care path that is most appropriate based on the symptoms and conditions of a patient.

Through the data collected, we were able to collect a useful vocabulary that covers medical terms to express symptoms and diseases that are prevalent in Malawi. We were able to collect over 1000 different expressions. This vocabulary can be expanded further. It can serve a good basis from which to develop a health vocabulary that is useful for improving communication but require further data collection and validation. Even in the case of Chichewa [11], which can be considered a major African language, no specialist dictionaries covering medical terminology for health have been developed. When available, interpreters often struggle with medical terminology. Our research provides evidence for the need, usefulness and feasibility of developing such a dictionary.

## Data Availability

The datasets used and/or analysed during the current study are available from the corresponding author upon reasonable request.

## 6. Limitations and further work

The findings of our study provide evidence attesting to language difficulties in the communication between healthcare professionals and patients in several facilities in Zomba. These challenges are significant and affect the quality of healthcare. This study can be expanded to cover more districts and healthcare facilities in Malawi. The extent of generalising our results to Malawi is influenced by the following limitations: the survey covers only one district of Malawi it is not a language study, so the vocabulary check we did is not sufficient for analysis the extent of the vocabulary needs of healthcare professionals or of that of patients. We collected data from 79 healthcare professionals and if the total numbers of healthcare professionals (112) have not significantly changed wince 2009, our sample represents 71% of the population.

Some contagion in responses was possible, several patients could have heard the responses given by others when translating terms in Chichewa for example. The diseases and symptoms mentioned by respondents may not reflect all the typical symptoms and conditions.

## 7. Conclusions

We studied language barriers in healthcare communication and provided evidence for the extent of language challenges faced by healthcare professionals and patients. This study is novel given the scarcity of linguistic research in Malawi in general, and in health communication in particular. We gathered evidence to attest that healthcare professionals use English throughout their training, for work reports and to record patient clinical history, but they face challenges in their communication with patients that must take place in local languages. The reality of urban and rural clinics is one of growing communication barriers caused by falling levels of education and literacy among patients, the inability of patients to express symptoms and to understand medical terms in their own language. Additionally, healthcare professionals also struggle to find Chichewa equivalent of medical terms when speaking to patients. Chichewa and English both act as additional languages for those with a different mother tongue, but their role in facilitating health communication is not perfect. The evidence we collected points to the need for developing medical resources such as dictionaries for both English and Chichewa that can facilitate translations and conversations about common medical conditions and symptoms. Additional studies on both language and cultural gap analysis are needed.

## 9. Supplementary material

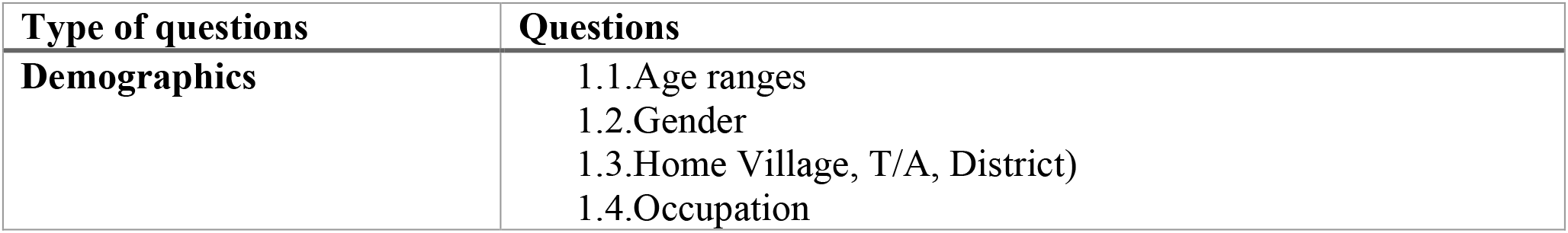
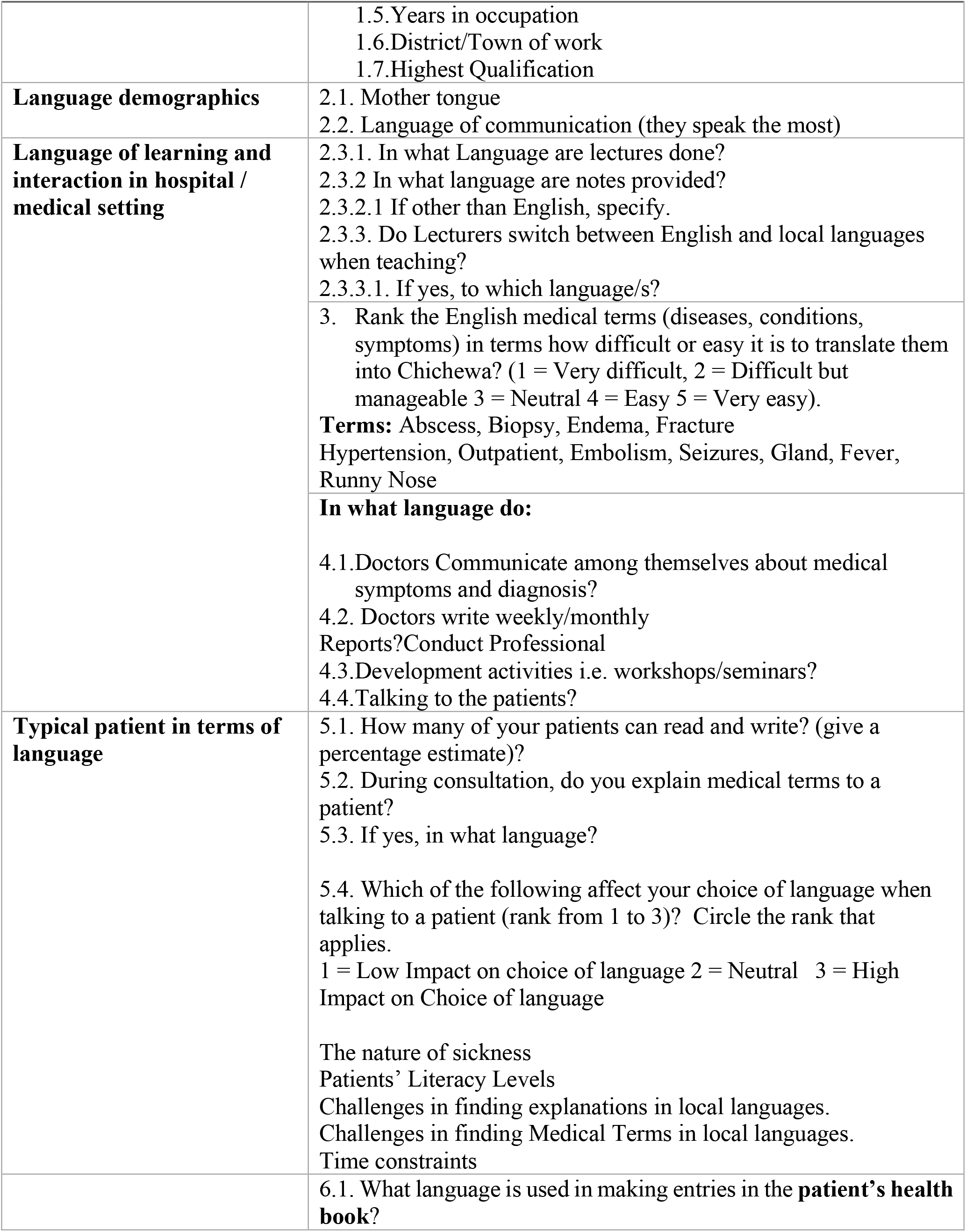
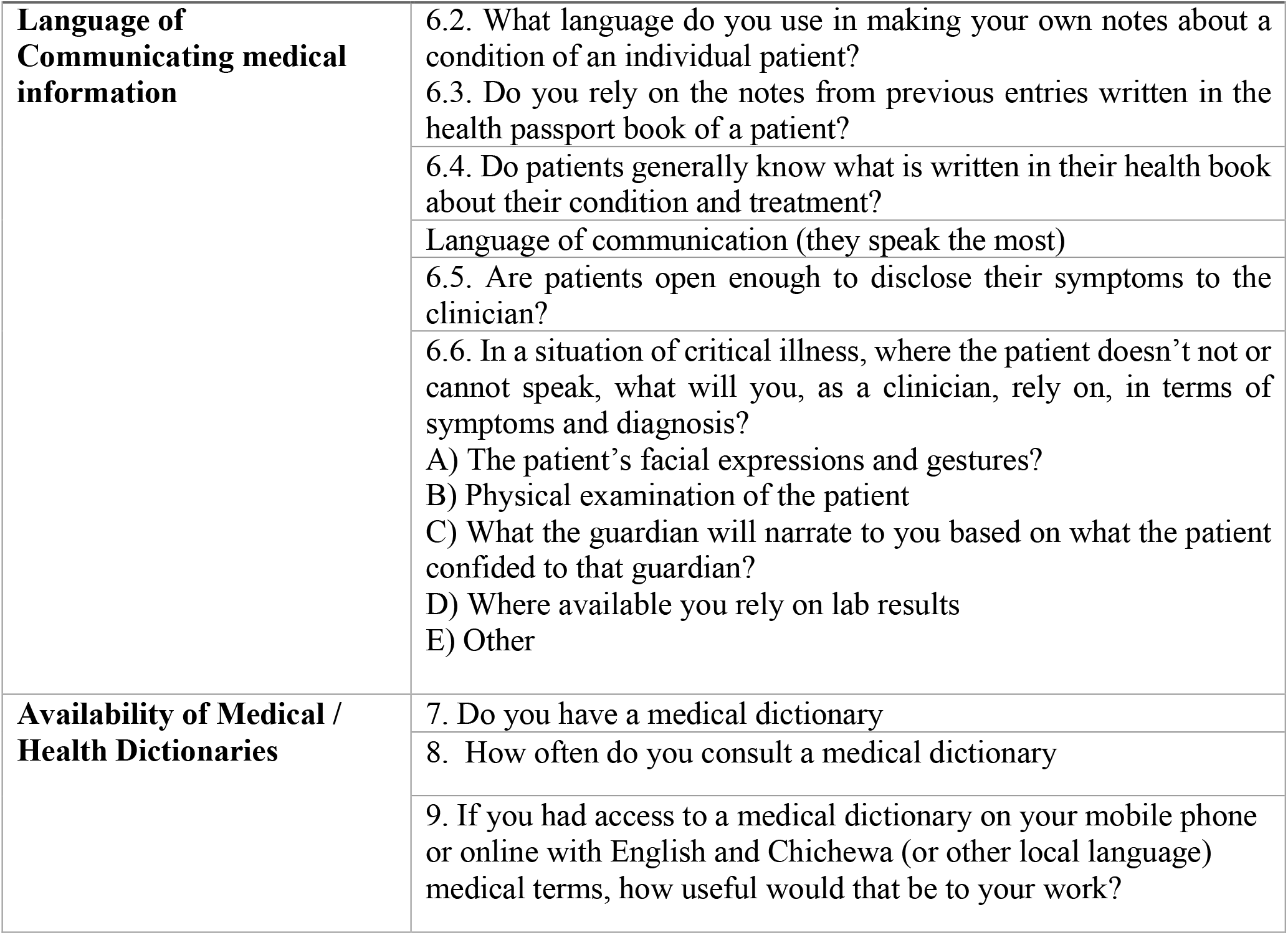

## Footnotes

1 UNESCO Institute for Statistics.

